# Pretrained Patient Trajectories for Adverse Drug Event Prediction Using Common Data Model-based Electronic Health Records

**DOI:** 10.1101/2024.09.30.24314595

**Authors:** Junmo Kim, Joo Seong Kim, Ji-Hyang Lee, Min-Gyu Kim, Taehyun Kim, Chaeeun Cho, Rae Woong Park, Kwangsoo Kim

## Abstract

**Background:** Pretraining electronic health record (EHR) data using language models by treating patient trajectories as natural language sentences has enhanced performance across various medical tasks. However, EHR pretraining models have never been utilized in adverse drug event (ADE) prediction.

**Methods:** A retrospective study was conducted on observational medical outcomes partnership (OMOP)-common data model (CDM) based EHR data from two separate tertiary hospitals. The data included patient information in various domains such as diagnosis, prescription, measurement, and procedure. For pretraining, codes were randomly masked, and the model was trained to infer the masked tokens utilizing preceding and following history. In this process, we introduced domain embedding (DE) to provide information about the domain of the masked token, preventing the model from finding codes from irrelevant domains. For qualitative analysis, we identified important features using the attention matrix from each finetuned model.

**Results:** 510,879 and 419,505 adult inpatients from two separate tertiary hospitals were included in internal and external datasets. EHR pretraining model with DE outperformed all the other baselines in all cohorts. For feature importance analysis, we demonstrated that the results were consistent with priorly reported background clinical knowledge. In addition to cohort-level interpretation, patient-level interpretation was also available.

**Conclusions:** CDM-based EHR pretraining model with DE is a proper model for various ADE prediction tasks. The results of the qualitative analysis with feature importance were consistent with background clinical knowledge.

**Plain language summary:** Patient history is like natural language; each medical code corresponds to a word, and the sequence of medical codes corresponds to a sentence. Language models learn from sentences by inferring masked words using remaining unmasked words, and language model-based EHR pretraining models comprehend medical context similarly. As several studies that have utilized EHR pretraining models have achieved great success in various tasks, we applied EHR pretraining models for adverse drug event (ADE) prediction. For better inference of the EHR pretraining model, we introduced domain embedding (DE) to provide a hint of the domain for each masked code. The model pretrained with DE performed the best in various ADE tasks, and regarding background clinical knowledge was well-reflected in our feature importance-based qualitative analysis.

## Introduction

Hospitals generate a lot of data every day, including diagnoses, measurements, prescriptions, procedures, and more, which are stored in electronic health records (EHR). The adoption of EHR has greatly increased in many countries. The rate of EHR adoption now exceeds 96%, 94%, 88%, and 96% in the US, UK, China, and South Korea (hereafter Korea), respectively.^1–3^ The widespread use of EHR has increased the amount of healthcare data, and numerous machine learning studies have been conducted using EHR data for various medical tasks, such as disease prediction, patient status monitoring, and diagnosis support.^4–6^

With the availability of large patient data, deep learning-based EHR analysis models have also been developed, and learning patient representation through pretraining has become an area ripe for exploration.^7–12^ As the pretraining-finetuning paradigm has achieved tremendous success in natural language processing, several studies proposed EHR pretraining models that learn patient representations from the sequential sets of medical codes corresponding to sentences in natural language. For pretraining tasks, most of those prior studies (e.g., BEHRT^9^, Med-BERT^10^, and CEHR-BERT^11^) utilized bidirectional encoder representation from transformers (BERT)^13^, one of the most famous models for contextualizing sequential inputs, especially natural language. As original BERT masks some of the tokens (words) from the sequential inputs (sentences) and infers the right tokens for the masked parts, BERT-based EHR pretraining models mask some of the medical codes and infer proper medical codes using unmasked preceding and following medical history. Prior studies achieved great performance in predicting several diseases, such as pancreatic cancer^10^, heart failure^11^, and non-accidental trauma^7^, when finetuned initially pretrained models compared to initially randomized (not pretrained) models.

EHR pretraining models exhibited great performance in various medical tasks, but adverse drug event (ADE) prediction using EHR pretraining models is a field much left to explore. Tertiary hospitals have good-quality inpatient records since all medical events that occur in the hospital are recorded in the EHR database. For instance, lab or vital signs variation before and after medication intake and condition and procedure as a coping strategy for ADE, which are stored in EHR, can reflect the topical or systemic impact of numerous drugs for various types of patients. Therefore, EHR provides new opportunities to address limitations of traditional approaches and promote discovery-oriented pharmacovigilance.^14^ Previous studies utilized odds ratio, regression-based modeling (e.g., logistic, Poisson, and Cox regression), and tree-based machine learning models for EHR-based ADE management, but no study had utilized EHR pretraining models for ADE prediction.

In this study, we exhibited the validity of EHR pretraining model in ADE prediction tasks using EHR data based on an observational medical outcomes partnership (OMOP)-common data model (CDM).^15,16^ OMOP-CDM (hereafter CDM) enables the transformation of different databases to a standardized format, facilitating multicenter study with different EHR databases and external validation of statistical models or artificial intelligence methodologies for EHR data.^17^ We used enormous records of diagnosis, prescription, measurement, and procedure domains in CDM from two separate tertiary hospitals in Korea. For pretraining, we adopted the masked language model (MLM). Codes were randomly masked, and EHR pretraining models were trained to infer the masked tokens with preceding and following history along with age and date. In this process, we introduced domain embedding (DE) to provide information about the domain of the masked token and prevent the model from finding unnecessary codes from other domains (Fig. 1a). EHR pretraining models with DE outperformed all the other baselines in several representative ADE prediction tasks. For the qualitative analysis, we identified important features at cohort and patient levels and demonstrated that the results were consistent with background clinical knowledge.

**Fig. 1.**
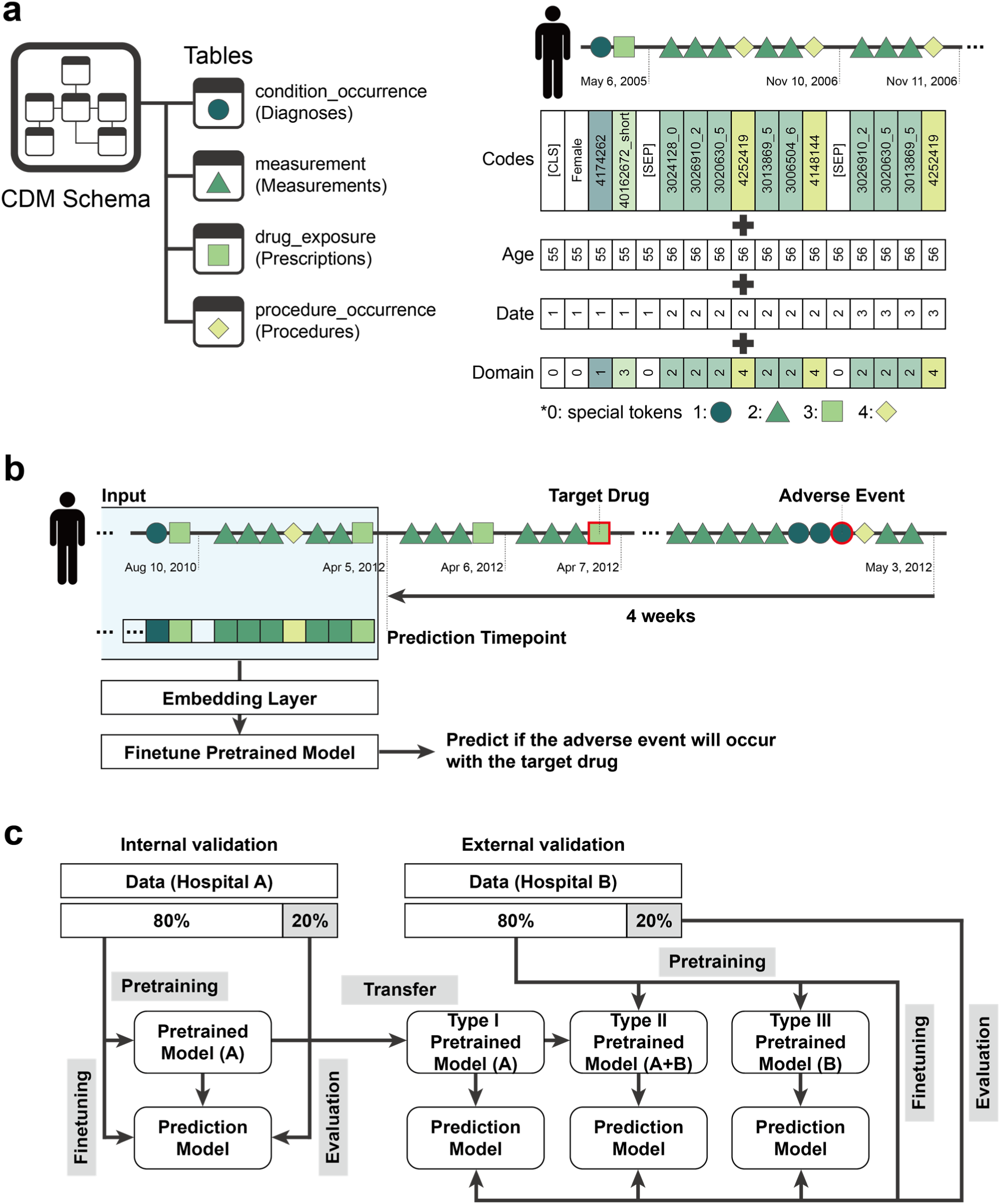
Workflow of ADE prediction using CDM-based EHR pretraining model. **a**, Data from four domains (diagnosis, measurement, prescription, and procedure) of OMOP-CDM were utilized. Each patient trajectory consists of medical records sorted by time, starts with [CLS] token and gender token, and are divided by [SEP] token for every day. Patient trajectory embedding is the summation of code, age, date, and domain embeddings.

Domain embedding has five tokens corresponding to special tokens and four domains of OMOP-CDM. **b**, For the case group, ADE occurred within four weeks after prescription, and the prediction timepoint was four weeks before the occurrence of adverse events. Each patient trajectory before the prediction timepoint was utilized to predict the occurrence of ADE. **c**, For internal validation, we finetuned the pretrained model to predict ADE and evaluated the prediction model. For external validation, we utilized three types of pretrained models: For type I, we transferred the model pretrained by the internal dataset and finetuned the model with the external dataset. For type II, to mitigate the gap in data distribution between two separate hospitals, we used the external dataset to additionally pretrain the model initially pretrained by the internal dataset. For type III, we pretrained the model only using the external dataset.

## Methods

### Data curation

This study used data from the Seoul National University Hospital (SNUH) between January 2001 and December 2023 and the Ajou University Medical Center (AUMC) between January 2004 and December 2023. Both hospitals operate EHR database based on OMOP-CDM version 5.3.^17^ We collected data from four domains (tables) of CDM schema, diagnosis (condition_occurrence), prescription (drug_exposure), measurement (measurement), and procedure (procedure_occurrence). Terminologies of diagnosis and procedure, measurement, and prescription are based on SNOMED CT^18^, Rx-Norm^19^, and LOINC^20^, respectively.

To screen patients with high-quality records, we included patients who had been hospitalized for at least three days in the study population, and patients under 18 were excluded. Data were randomly split into training (70%), validation (10%), and hold-out test (20%) datasets (Fig. 1c).

The Institutional Review Board (IRB) of Seoul National University Hospital (IRB approval No. 2406-060-1543) approved the study with a waiver of informed consent, considering that our study used retrospective and observational EHR data. The approval aligns with the principles outlined in the Declaration of Helsinki, the Korean Bioethics and Safety Act (Law No. 16372), and the Human Research Protection Program–Standard Operating Procedure of Seoul National University Hospital.

### ADE prediction task

For drug groups, we included nonsteroidal anti-inflammatory drugs (NSAID), anticoagulants (AC), and chemotherapy (Chemo). For corresponding adverse events, we selected peptic ulcer (PU), intracranial hemorrhage (ICH), and neutropenic fever (NF), respectively. The information on all drugs included in each drug group is summarized in Supplementary Table 1, and all concept IDs for each adverse event are summarized in Supplementary Table 2. For each cohort, we first included patients who were prescribed the target drug. Those who were diagnosed with corresponding adverse events within four weeks after the prescription were included in the case group (Fig. 1b), and those who had no record of adverse events were included in the control group. For the case group, we set a prediction timepoint to four weeks before the adverse event occurrence to prevent target leakage, assuming that no obvious symptom records directly related to the target ADE exist prior to this timepoint. For the control group, we randomly selected one of the days on which the target drug was prescribed and set it as the prediction timepoint to minimize potential bias caused by selection criteria. Given that the duration of action can vary for different drugs and circumstances, we conducted additional experiments by setting the prediction timepoints to two, eight, and twelve weeks before the occurrence of adverse events. In addition, diverse prediction timepoints were to consider impact of the different amount of information.

### Record tokenization

Each diagnosis or procedure record was tokenized by its corresponding concept ID in SNOMED-CT. For each prescription, we added ‘short’ at the end of the corresponding concept ID in Rx-Norm if its duration was shorter than four weeks and ‘long’ if it was more than four weeks (Fig. 1a). This process was to distinguish prescription type, considering inpatients are usually prescribed daily (or with every meal), while outpatients typically receive prescriptions for longer periods. Each measurement item was categorized into ten tokens divided into deciles, and we added a corresponding decile number between 0 and 9 at the end of its corresponding concept ID in LOINC (Fig. 1a). For example, in Fig. 1a, ‘40162672_short’ indicates prescription of amitriptyline hydrochloride 10mg oral tablet with less than four weeks, and ‘3026910_2’ indicates low level (between 2^nd^ and 3^rd^ decile) of gamma-glutamyl transferase (GGT). We only utilized numerical measurement items because the items with natural language results were unstructured and unformatted to tokenize. Regarding special tokens, we utilized five special tokens: [PAD], [MASK], [UNK], [CLS], and [SEP].^13^ [PAD] was used to align patient trajectories with different lengths to the same lengths, and [MASK] was used for the MLM process. Tokens that were not in the training dataset were replaced with [UNK]. Every trajectory starts with [CLS] considering representation usage for various downstream tasks. Like BERT used [SEP] to separate two sentences, we inserted [SEP] between all different dates. The token counts for each domain in each dataset are summarized in Supplementary Table 3. Ages were simply inserted as integer tokens, and dates were grouped by integer tokens increasing by one. We used five tokens for domains: special tokens, diagnosis, measurement, prescription, and procedure (Fig. 1a).

### Trajectory construction and embedding

All tokens were sorted in order of time for each patient, and the maximum trajectory length was set to 2048, covering more than 85% of all trajectories. All trajectories start with [CLS] and gender tokens (Fig. 1a). For pretraining, trajectories longer than the maximum length were sliced into non-overlapping sub-trajectories with the maximum length to prevent potential dependency among trajectories from a single patient. For finetuning, we first removed tokens after the prediction timepoint, and then used the latest 2048 tokens instead of slicing them into sub-trajectories. Trajectories with less than 50 tokens were excluded because predicting ADE in patients with too little information was not considered to be worthwhile. All short trajectories were padded to a length of 2048, aligning them with the same length for mini-batch training. For each token embedding, corresponding embeddings of age, date, and domain tokens were added.

### Pretraining process

EHR pretraining models were based on BERT^10,13^ architecture, with six layers, eight attention heads, and a hidden dimension of 256. We aggregated the vocabulary sets from the internal and external datasets, and the final vocabulary size was 41,536. However, we set the embedding size of vocabulary to be 50,000, considering future additional medical codes. For age, date, and domain embedding, we set the embedding size to 180, 1024, and 20, respectively. The size of domain embedding was decided considering additional domains like unstructured data (e.g., X-ray, electrocardiogram, and nursing report). For pretraining, we randomly masked 30% of tokens except special tokens for each trajectory, and the model was trained to infer the masked tokens of specific domains with preceding and following history along with age, date, and domain. The models were pretrained with 100 epochs, and we selected the model with the minimum CE loss during the training.

For external validation, we introduced three types of EHR pretraining model (Fig. 1c): For type I, we used the model initially pretrained with the internal dataset without additional pretraining. For type II, we additionally pretrained the model that was initially pretrained by the internal dataset using the external dataset. For type III, we initially pretrained the model only using the external dataset. The type II and III models were pretrained using the external dataset for ten epochs.

### Finetuning process

For finetuning for ADE prediction, we added feed-forward neural network (FFNN), hyperbolic tangent (Tanh) activation function, dropout, and FFNN on the representation of the first token ([CLS]) to yield a vector of size two for binary classification. Finetuning was trained to minimize binary CE loss for ADE prediction. We recorded the classification performance for every cycle of 100 batches, and the finetuning was stopped if the area under the receiver operating characteristic curve (AUROC) of the validation dataset did not improve for ten cycles (1000 batches). Considering extreme class imbalance and small batch size, we randomly oversampled the minority class to be one-tenth of the majority class during finetuning.

### Feature importance

We calculated the feature importance using attention matrices of case groups. We averaged attention matrices of eight attention heads from the last self-attention layer to yield a final attention matrix (hereafter attention matrix). An attention matrix A for each trajectory was 2048 (maximum trajectory length) by 2048 matrix, and an attention score A_j_ indicates how closely i^th^ and j^th^ tokens are related.^21^ Note that ∑_j_ *A_ij_* = 1 ∀i ∈ (0, …, 2048).

For comprehensive analysis, we first averaged the values of the same tokens in each row of the attention matrix to prevent repeatedly appearing tokens of routine medical events (e.g., normal saline, blood pressure, and electrocardiogram) from getting unnecessarily high scores. Then, we extracted the maximum value among 2048 rows of each token. Finally, each 2048 by 2048 attention matrix was transformed into a one-dimensional vector. We called this vector as trajectory attention vector (TAV). The value of each element of TAV is as follows:

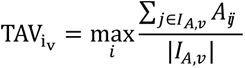

where *i_v_* is the index of code *v* in TAV and *I_A,v_* is a set of indices of code *v* in the original attention matrix *A*. TAV finally indicates the importance of each token in the trajectory. In each TAV, we excluded tokens that appeared in fewer than 5% of the patients in the case group. This process was to prevent irrelevant trivial tokens coincidently having high attention scores from becoming important features. Finally, we averaged all TAV values by tokens and selected the top 10 most important tokens from each domain.

For individualized analysis, we simply summed all rows of an attention matrix and then extracted the top 10% of tokens with the highest values. This process was to enhance readability because most trajectories had hundreds of tokens.

### Statistical analysis

Characteristics (age, sex, and comorbidities) in internal and external cohorts were compared by calculating P-values using the Student’s t-test for age and the Fisher’s exact test for the rest of the variables. To measure and compare the performances of the models, we mainly used AUROC. We also provided area under the precision-recall curve (AUPRC), sensitivity, specificity, precision, and F1-score. Confidence intervals of AUROC and AUPRC were calculated by DeLong’s method^22^, while those of F1-score, sensitivity, specificity, and precision were calculated by Wilson’s method^23^. Statistical significance was set at α=0.05. All statistical analyses were performed using scikit-learn (version 1.0.2) in Python (version 3.8.10).

### Training and computing information

The learning rate and dropout rate were set to 5e-5 and 0.1, and the batch size was set to 16. For deep learning and BERT implementation, we used Pytorch (version 1.12.0) and HuggingFace package (version 4.41.2)^24^ in Python (version 3.8.10). We used four NVIDIA RTX A6000 GPUs for internal processes and four NVIDIA GeForce RTX 3090 Ti GPUs for external processes.

## Results

### Study population

We used records of 510,879 and 419,505 adult patients (aged over 18) who were hospitalized for at least three days in SNUH and AUMC, respectively. The baseline characteristics of the datasets are summarized in Table 1. The patients in AUMC had more codes than SNUH, while the median days of hospitalization were around a month for both hospitals. The distribution of comorbidities was different between the two hospitals: The rates of dementia, renal disease, and malignant tumor were higher in the internal dataset, while the rest of the comorbidities were higher in the external dataset.

**Table 1.**
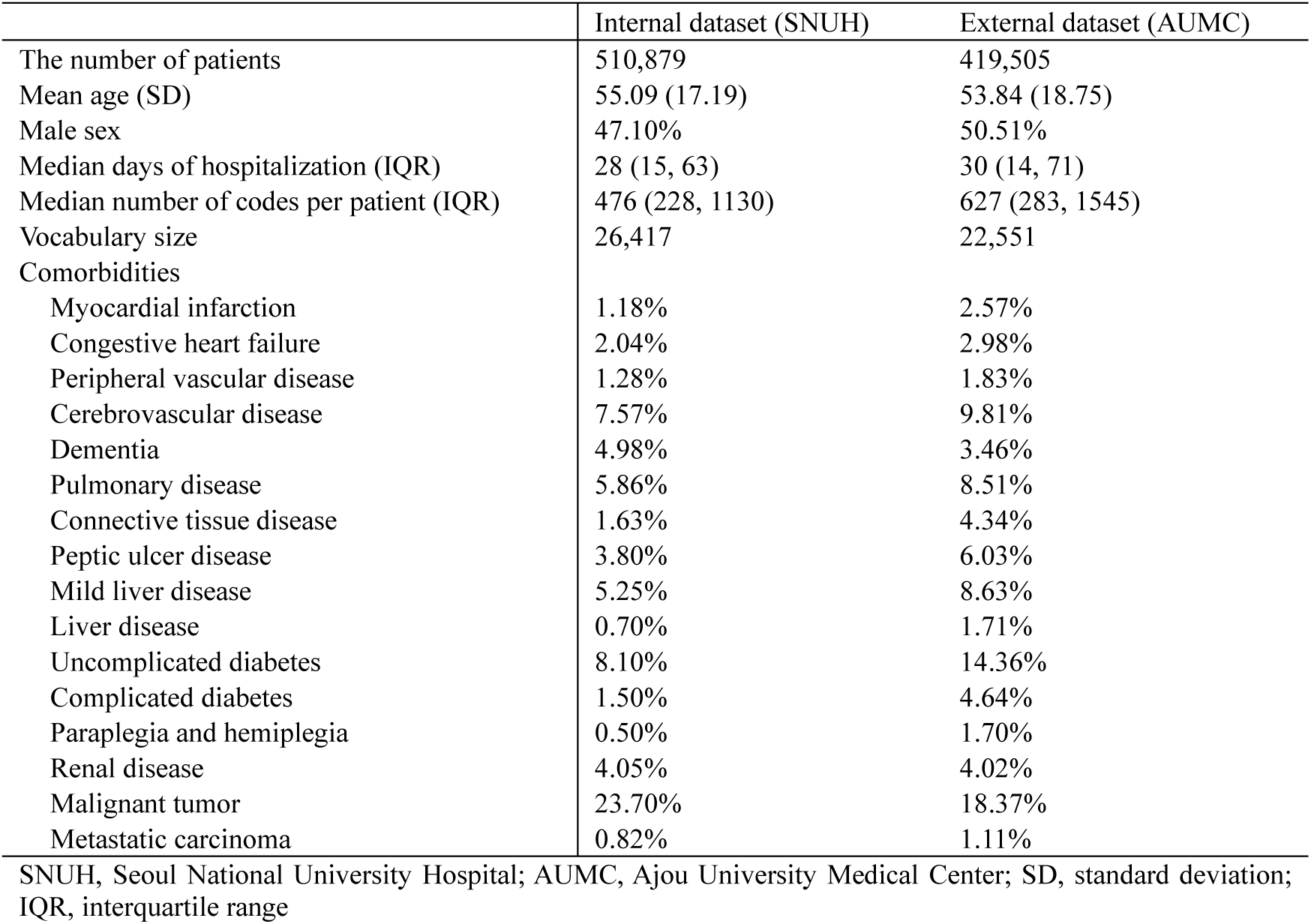
Baseline characteristics of datasets.

|  | Internal dataset (SNUH) | External dataset (AUMC) |
| --- | --- | --- |
| The number of patients | 510,879 | 419,505 |
| Mean age (SD) | 55.09 (17.19) | 53.84 (18.75) |
| Male sex | 47.10% | 50.51% |
| Median days of hospitalization (IQR) | 28 (15, 63) | 30 (14, 71) |
| Median number of codes per patient (IQR) | 476 (228, 1130) | 627 (283, 1545) |
| Vocabulary size | 26,417 | 22,551 |
| Comorbidities |  |  |
| Myocardial infarction | 1.18% | 2.57% |
| Congestive heart failure | 2.04% | 2.98% |
| Peripheral vascular disease | 1.28% | 1.83% |
| Cerebrovascular disease | 7.57% | 9.81% |
| Dementia | 4.98% | 3.46% |
| Pulmonary disease | 5.86% | 8.51% |
| Connective tissue disease | 1.63% | 4.34% |
| Peptic ulcer disease | 3.80% | 6.03% |
| Mild liver disease | 5.25% | 8.63% |
| Liver disease | 0.70% | 1.71% |
| Uncomplicated diabetes | 8.10% | 14.36% |
| Complicated diabetes | 1.50% | 4.64% |
| Paraplegia and hemiplegia | 0.50% | 1.70% |
| Renal disease | 4.05% | 4.02% |
| Malignant tumor | 23.70% | 18.37% |
| Metastatic carcinoma | 0.82% | 1.11% |

SNUH, Seoul National University Hospital; AUMC, Ajou University Medical Center; SD, standard deviation; IQR, interquartile range

Patient demographics of each cohort from both hospitals are summarized in Supplementary Table 4. As the prediction timepoint moved back to the past and the monitoring period for adverse events increased, the number of patients in each case group also increased in NSAID-PU and AC-ICH cohorts. For the Chemo-NF cohort, more patients were excluded due to a lack of previous records than those included by an increased monitoring period.

### Pretraining results

The validation losses of MLMs were much lower with DE (Supplementary Fig. 1). This result indicates that finding the correct codes for masked tokens was much easier with a hint of domain. For pretraining external validation, the validation loss of type II pretrained models was much lower than the type III, even though those models were pretrained by the same external dataset. In addition, even with a few epochs, the validation loss of the type II models showed convergence (Supplementary Fig. 1b).

### Performance evaluation

The model pretrained with DE outperformed all the other models in all metrics in all cohorts (Table 2). The pretraining process improved AUROC and AUPRC for all cohorts (pretrained without DE), but the adoption of DE improved the performance much more. When the model was initialized with randomized weights, the adoption of DE hardly influenced the performance.

**Table 2.** Internal validation of finetuned models for all cohorts.

|  | AUROC | AUPRC | Sensitivity | Specificity | Precision | F1-score |
| --- | --- | --- | --- | --- | --- | --- |
| NSAID (PU) |  |  |  |  |  |  |
| Randomized without DE | 0.848<br>(0.833-0.864) | 0.044<br>(0.029-0.060) | 0.836<br>(0.832-0.839) | 0.727<br>(0.724-0.731) | 0.025<br>(0.024-0.027) | 0.049<br>(0.047-0.051) |
| Randomized with DE | 0.848<br>(0.833-0.864) | 0.044<br>(0.029-0.060) | 0.836<br>(0.832-0.839) | 0.727<br>(0.723-0.731) | 0.025<br>(0.024-0.027) | 0.049<br>(0.047-0.051) |
| Pretrained without DE | 0.954<br>(0.946-0.962) | 0.357<br>(0.349-0.365) | 0.909<br>(0.906-0.911) | 0.870<br>(0.867-0.873) | 0.056<br>(0.054-0.058) | 0.105<br>(0.103-0.108) |
| Pretrained with DE | **0.977**<br>**(0.971-0.984)** | **0.668**<br>**(0.661-0.675)** | **0.942**<br>**(0.940-0.944)** | **0.916**<br>**(0.913-0.918)** | **0.087**<br>**(0.084-0.089)** | **0.159**<br>**(0.156-0.162)** |
| AC (ICH) |  |  |  |  |  |  |
| Randomized without DE | 0.769<br>(0.739-0.799) | 0.022<br>(0.000-0.799) | 0.753<br>(0.749-0.758) | 0.683<br>(0.678-0.688) | 0.016<br>(0.015-0.018) | 0.032<br>(0.030-0.034) |
| Randomized with DE | 0.769<br>(0.739-0.799) | 0.022<br>(0.000-0.799) | 0.753<br>(0.749-0.758) | 0.683<br>(0.678-0.688) | 0.016<br>(0.015-0.018) | 0.032<br>(0.030-0.034) |
| Pretrained without DE | 0.822<br>(0.797-0.848) | 0.040<br>(0.015-0.065) | 0.836<br>(0.831-0.840) | 0.670<br>(0.665-0.675) | 0.017<br>(0.016-0.019) | 0.034<br>(0.032-0.036) |
| Pretrained with DE | **0.908**<br>**(0.886-0.931)** | **0.206**<br>**(0.183-0.228)** | **0.840**<br>**(0.836-0.844)** | **0.863**<br>**(0.859-0.867)** | **0.041**<br>**(0.039-0.043)** | **0.078**<br>**(0.075-0.081)** |
| Chemo (NF) |  |  |  |  |  |  |
| Randomized without DE | 0.833<br>(0.817-0.848) | 0.140<br>(0.124-0.156) | 0.867<br>(0.862-0.872) | 0.669<br>(0.662-0.676) | 0.072<br>(0.068-0.076) | 0.132<br>(0.127-0.137) |
| Randomized with DE | 0.833<br>(0.817-0.849) | 0.140<br>(0.124-0.156) | 0.867<br>(0.862-0.872) | 0.668<br>(0.661-0.675) | 0.071<br>(0.068-0.075) | 0.132<br>(0.127-0.137) |
| Pretrained without DE | 0.971<br>(0.966-0.976) | 0.704<br>(0.698-0.709) | 0.933<br>(0.929-0.936) | 0.890<br>(0.885-0.895) | 0.200<br>(0.194-0.206) | 0.329<br>(0.322-0.337) |
| Pretrained with DE | **0.989**<br>**(0.985-0.993)** | **0.909**<br>**(0.905-0.912)** | **0.953**<br>**(0.950-0.956)** | **0.948**<br>**(0.945-0.951)** | **0.350**<br>**(0.343-0.358)** | **0.512**<br>**(0.505-0.520)** |

Bold indicates the best. Sensitivity, specificity, precision, and F1-score were calculated by Youden's index. Confidence intervals (CIs) of AUROC and AUPRC were calculated by DeLong's method. CIs of sensitivity, specificity, precision, and F1-score were calculated by Wilson's method. NSAID, nonsteroidal anti-inflammatory drugs; PU, peptic ulcer; AC, anticoagulants; ICH, intracranial hemorrhage; Chemo, chemotherapy; NF, neutropenic fever; AUROC, area under the receiver operating characteristic curve; AUPRC, area under the precision-recall curve

For external validation, the adoption of DE improved AUROC and AUPRC of the type I and II models in all cohorts (Table 3). For the type I with DE, even though the model was pretrained from the different hospital data (SNUH) and there was no additional pretraining, the AUROC was much higher than the randomized model in all cohorts (AUROC from 0.814 to 0.947 in NSAID-PU cohort, from 0.769 to 0.940 in AC-ICH cohort, and from 0.918 to 0.934 in Chemo-NF cohort). For the type II and III, the type II exhibited higher AUROC and AUPRC in all cohorts. Considering those two models were pretrained for the same ten epochs, the model initially pretrained by the other dataset (type II) was more effective than the initially randomized model (type III). Like internal validation results, the adoption of DE had minimal effect on performance in initially randomized models. The receiver operating characteristic (ROC), precision-recall (PR), and calibration curves of internal and external validations are summarized in Fig. 2. Since the proportion of patients with adverse events was much less than those without (Supplementary Table 3), most models overpredicted the risk of adverse events.

**Table 3.** External validation of finetuned models for all cohorts.

|  | AUROC | AUPRC | Sensitivity | Specificity | Precision | F1-score |
| --- | --- | --- | --- | --- | --- | --- |
| NSAID (PU) |  |  |  |  |  |  |
| Randomized without DE | 0.814<br>(0.798-0.830) | 0.070<br>(0.054-0.086) | 0.804<br>(0.800-0.808) | 0.691<br>(0.686-0.696) | 0.042<br>(0.040-0.044) | 0.079<br>(0.076-0.082) |
| Randomized with DE | 0.814<br>(0.798-0.830) | 0.070<br>(0.054-0.086) | 0.804<br>(0.800-0.808) | 0.690<br>(0.685-0.695) | 0.041<br>(0.039-0.044) | 0.079<br>(0.076-0.082) |
| Pretrained without DE (Type I) | 0.834<br>(0.819-0.848) | 0.077<br>(0.063-0.092) | 0.793<br>(0.788-0.797) | 0.724<br>(0.719-0.729) | 0.046<br>(0.044-0.048) | 0.086<br>(0.083-0.089) |
| Pretrained with DE (Type I) | 0.947<br>(0.938-0.956) | 0.575<br>(0.566-0.584) | 0.847<br>(0.843-0.851) | 0.904<br>(0.901-0.907) | 0.128<br>(0.124-0.131) | 0.222<br>(0.218-0.226) |
| Pretrained without DE (Type II) | 0.919<br>(0.908-0.929) | 0.278<br>(0.268-0.289) | 0.867<br>(0.864-0.871) | 0.826<br>(0.822-0.830) | 0.077<br>(0.074-0.079) | 0.141<br>(0.137-0.144) |
| Pretrained with DE (Type II) | **0.971**<br>**(0.964-0.978)** | **0.712**<br>**(0.705-0.719)** | **0.908**<br>**(0.905-0.911)** | **0.930**<br>**(0.927-0.933)** | **0.178**<br>**(0.174-0.182)** | **0.297**<br>**(0.292-0.302)** |
| Pretrained without DE (Type III) | 0.857<br>(0.844-0.871) | 0.118<br>(0.104-0.132) | 0.883<br>(0.879-0.886) | 0.679<br>(0.674-0.684) | 0.044<br>(0.042-0.046) | 0.083<br>(0.081-0.086) |
| Pretrained with DE (Type III) | 0.925<br>(0.913-0.936) | 0.435<br>(0.423-0.446) | 0.857<br>(0.853-0.861) | 0.839<br>(0.836-0.843) | 0.082<br>(0.079-0.084) | 0.149<br>(0.145-0.153) |
| AC (ICH) |  |  |  |  |  |  |
| Randomized without DE | 0.769<br>(0.738-0.799) | 0.043<br>(0.013-0.074) | 0.735<br>(0.729-0.741) | 0.678<br>(0.672-0.684) | 0.022<br>(0.020-0.024) | 0.043<br>(0.040-0.045) |
| Randomized with DE | 0.769<br>(0.738-0.799) | 0.044<br>(0.013-0.074) | 0.735<br>(0.729-0.741) | 0.678<br>(0.672-0.683) | 0.022<br>(0.020-0.024) | 0.043<br>(0.040-0.045) |
| Pretrained without DE (Type I) | 0.856<br>(0.831-0.881) | 0.265<br>(0.240-0.290) | 0.774<br>(0.768-0.779) | 0.790<br>(0.785-0.795) | 0.035<br>(0.033-0.037) | 0.067<br>(0.064-0.070) |
| Pretrained with DE (Type I) | 0.940<br>(0.923-0.958) | 0.570<br>(0.552-0.588) | 0.855<br>(0.850-0.859) | 0.891<br>(0.887-0.895) | 0.072<br>(0.068-0.075) | 0.132<br>(0.128-0.137) |
| Pretrained without DE (Type II) | 0.943<br>(0.927-0.959) | 0.573<br>(0.557-0.589) | 0.838<br>(0.833-0.842) | **0.900**<br>**(0.897-0.904)** | **0.076**<br>**(0.073-0.080)** | **0.140**<br>**(0.136-0.144)** |
| Pretrained with DE (Type II) | **0.972**<br>**(0.963-0.981)** | **0.673**<br>**(0.664-0.681)** | **0.957**<br>**(0.955-0.960)** | 0.853<br>(0.849-0.858) | 0.060<br>(0.057-0.063) | 0.113<br>(0.109-0.117) |
| Pretrained without DE (Type III) | 0.921<br>(0.902-0.940) | 0.469<br>(0.450-0.487) | 0.838<br>(0.833-0.842) | 0.865<br>(0.861-0.870) | 0.058<br>(0.055-0.061) | 0.108<br>(0.104-0.112) |
| Pretrained with DE (Type III) | 0.909<br>(0.887-0.930) | 0.532<br>(0.511-0.554) | 0.791<br>(0.785-0.796) | 0.869<br>(0.864-0.873) | 0.056<br>(0.053-0.059) | 0.104<br>(0.101-0.108) |
| Chemo (NF) |  |  |  |  |  |  |
| Randomized without DE | 0.918<br>(0.893-0.942) | 0.236<br>(0.212-0.261) | 0.892<br>(0.884-0.899) | 0.805<br>(0.795-0.814) | 0.053<br>(0.048-0.058) | 0.099<br>(0.093-0.107) |
| Randomized with DE | 0.918<br>(0.893-0.942) | 0.236<br>(0.212-0.261) | 0.892<br>(0.884-0.899) | 0.805<br>(0.795-0.814) | 0.053<br>(0.048-0.058) | 0.099<br>(0.093-0.107) |
| Pretrained without DE (Type I) | 0.909<br>(0.885-0.933) | 0.117<br>(0.093-0.141) | 0.904<br>(0.896-0.910) | 0.788<br>(0.779-0.798) | 0.049<br>(0.045-0.055) | 0.094<br>(0.087-0.101) |
| Pretrained with DE (Type I) | 0.934<br>(0.915-0.953) | 0.210<br>(0.191-0.229) | 0.928<br>(0.921-0.934) | 0.832<br>(0.823-0.841) | 0.063<br>(0.058-0.069) | 0.118<br>(0.111-0.126) |
| Pretrained without DE (Type II) | 0.938<br>(0.921-0.955) | 0.237<br>(0.220-0.254) | 0.880<br>(0.872-0.887) | 0.864<br>(0.855-0.872) | **0.073**<br>**(0.067-0.079)** | **0.135**<br>**(0.127-0.143)** |
| Pretrained with DE (Type II) | **0.951**<br>**(0.937-0.966)** | **0.368**<br>**(0.354-0.383)** | **0.988**<br>**(0.985-0.990)** | 0.768<br>(0.758-0.778) | 0.049<br>(0.044-0.055) | 0.094<br>(0.087-0.101) |
| Pretrained without DE (Type III) | 0.933<br>(0.910-0.955) | 0.278<br>(0.255-0.300) | 0.940<br>(0.934-0.945) | 0.798<br>(0.789-0.808) | 0.054<br>(0.049-0.059) | 0.102<br>(0.095-0.109) |
| Pretrained with DE (Type III) | 0.925<br>(0.902-0.948) | 0.231<br>(0.208-0.254) | 0.831<br>(0.822-0.840) | **0.867**<br>**(0.859-0.875)** | 0.071<br>(0.065-0.077) | 0.130<br>(0.123-0.138) |

Bold indicates the best. Sensitivity, specificity, precision, and F1-score were calculated using Youden's index. Confidence intervals (CIs) of AUROC and AUPRC were calculated using DeLong's method. CIs of sensitivity, specificity, precision, and F1-score were calculated using Wilson's method. Type I indicates the model initially pretrained by the internal dataset. Type II indicates the model that was initially pretrained by the internal dataset and was additionally pretrained by the external dataset. Type III indicates the model that was pretrained only by the external dataset. NSAID, nonsteroidal anti-inflammatory drug; PU, peptic ulcer; AC, anticoagulant; ICH, intracranial hemorrhage; Chemo, chemotherapy; NF, neutropenic fever; AUROC, area under the receiver operating characteristic curve; AUPRC, area under the precision-recall curve

**Fig. 2.**
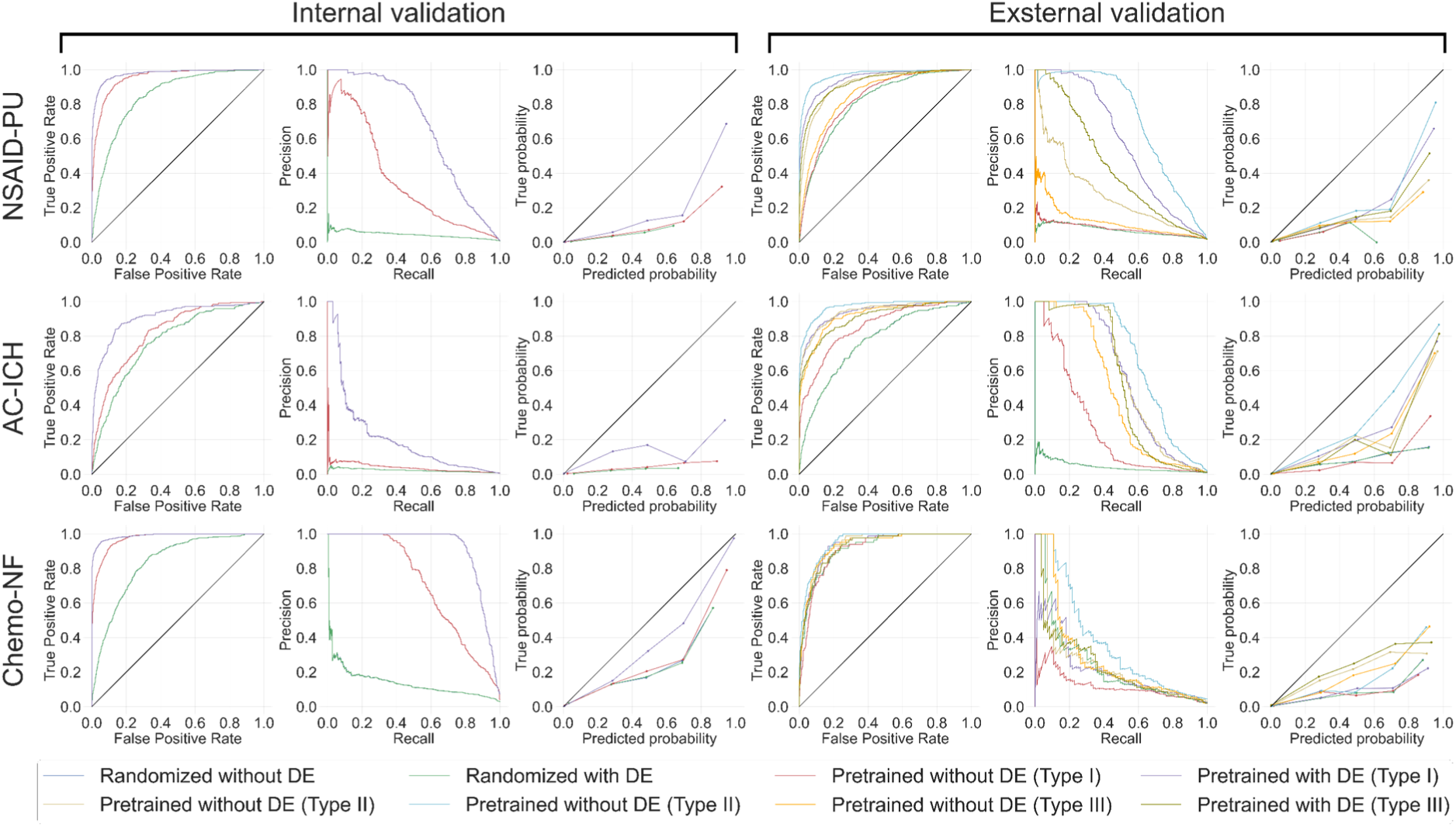
Receiver operating characteristic (ROC), precision-recall (PR), and calibration curves of internal and external validations. The left three columns are the results of internal validation and the right three columns are the results of external validation. The prediction cutoff point of each finetuned model was set at Youden’s index, which maximizes the sum of sensitivity (true positive rate) and specificity (1 - false positive rate). For both internal and external validation, most of the blue lines (for models randomized without DE) are invisible because they were overlapped by the green lines (for models randomized with DE).

### Analysis of different prediction timepoints

We set the prediction timepoint to four weeks before the occurrence of adverse events. However, since adverse events can occur at different times depending on the drug and circumstances, we conducted additional experiments by setting the prediction timepoints to two, eight, and twelve weeks before the occurrence of adverse events. For external validation, we utilized the type II pretrained model with DE because it was the best-performing type with a prediction timepoint of four weeks before ADE occurrence (Table 3). The model pretrained with DE exhibited the best AUROC and AUPRC in all cohorts with all timepoints, only except the Chemo-NF cohort of the external dataset with the prediction timepoint of twelve weeks (Supplementary Tables 5-10). Adoption of DE was effective in most cases with different prediction timepoints.

### Model interpretation

We reported the top 10 most important features from each domain, and various features relevant to each drug and adverse event were included in the top 10 (Figs. 3 and 4). In this process, we used the results of the model pretrained with DE (type II for external validation). For the NSAID-PU cohort in the internal validation, the model mainly focused on NSAIDs and aspirin prescription patterns for predicting PU. Celecoxib was the most important, even though it is known to be associated with a lower risk of gastrointestinal adverse events (such as PU).^25,26^ This might be due to patients switching to celecoxib after experiencing gastrointestinal symptoms while taking other NSAIDs; indeed, patients who took celecoxib were prescribed an average of 3.1 NSAIDs, while those not taking celecoxib were prescribed an average of 1.6 NSAIDs. Clopidogrel is usually prescribed with aspirin for the prevention of cardiovascular events.^27^ Clopidogrel can be associated with several diagnoses and procedures regarding cardiovascular diseases (angina pectoris, hypertension, cerebral infarction, and all of the included procedures). Gastric symptoms such as chest pain and nonulcer dyspepsia were also important. In the external validation, similarly, the model paid attention to NSAIDs and aspirin prescription patterns. For diagnosis, several conditions that are usually treated by NSAIDs (headache, joint pain, and pain caused by periodontitis) were important, and like the internal validation, chest pain was one of the main factors. Headache can be associated with another important feature, cerebral infarction. Valid medical context existed; however, more investigation is needed into the reason why cardiovascular diseases were important in the NSAID-PU cohort.

**Fig. 3.**
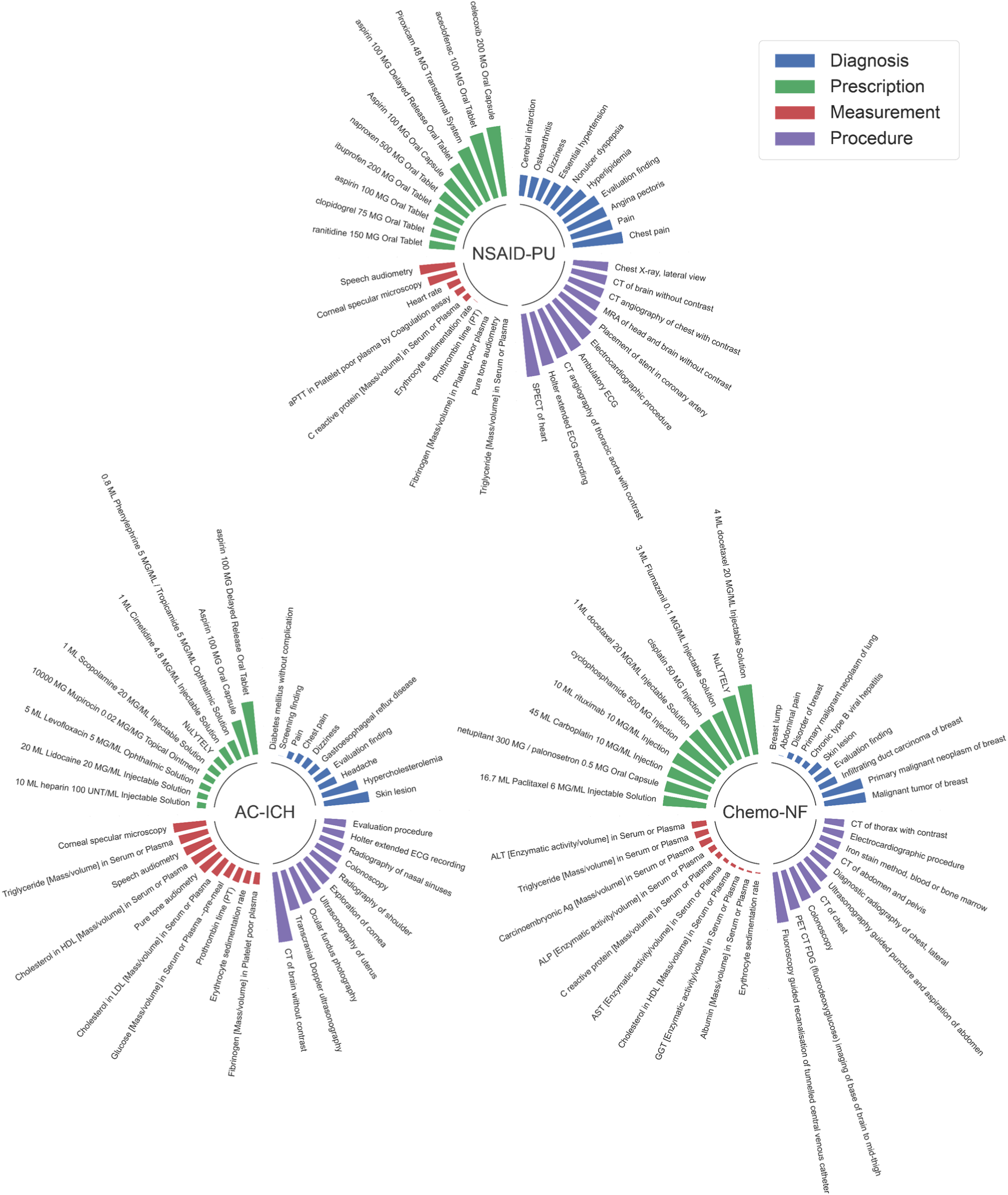
Feature importance of models finetuned with the internal dataset. Top 10 important features in each domain. Feature importance was deduced based on attention scores in the model. To prevent routine medical events (e.g., normal saline, blood pressure, and electrocardiogram) that repeatedly appear in trajectories to be excessively important, attention scores of the same tokens were initially averaged at the trajectory level, and then the scores of each token were finally averaged. For each cohort, the length of the longest bar was fixed to be the same, and the lengths of all the rest of the features were normalized accordingly.

**Fig. 4.**
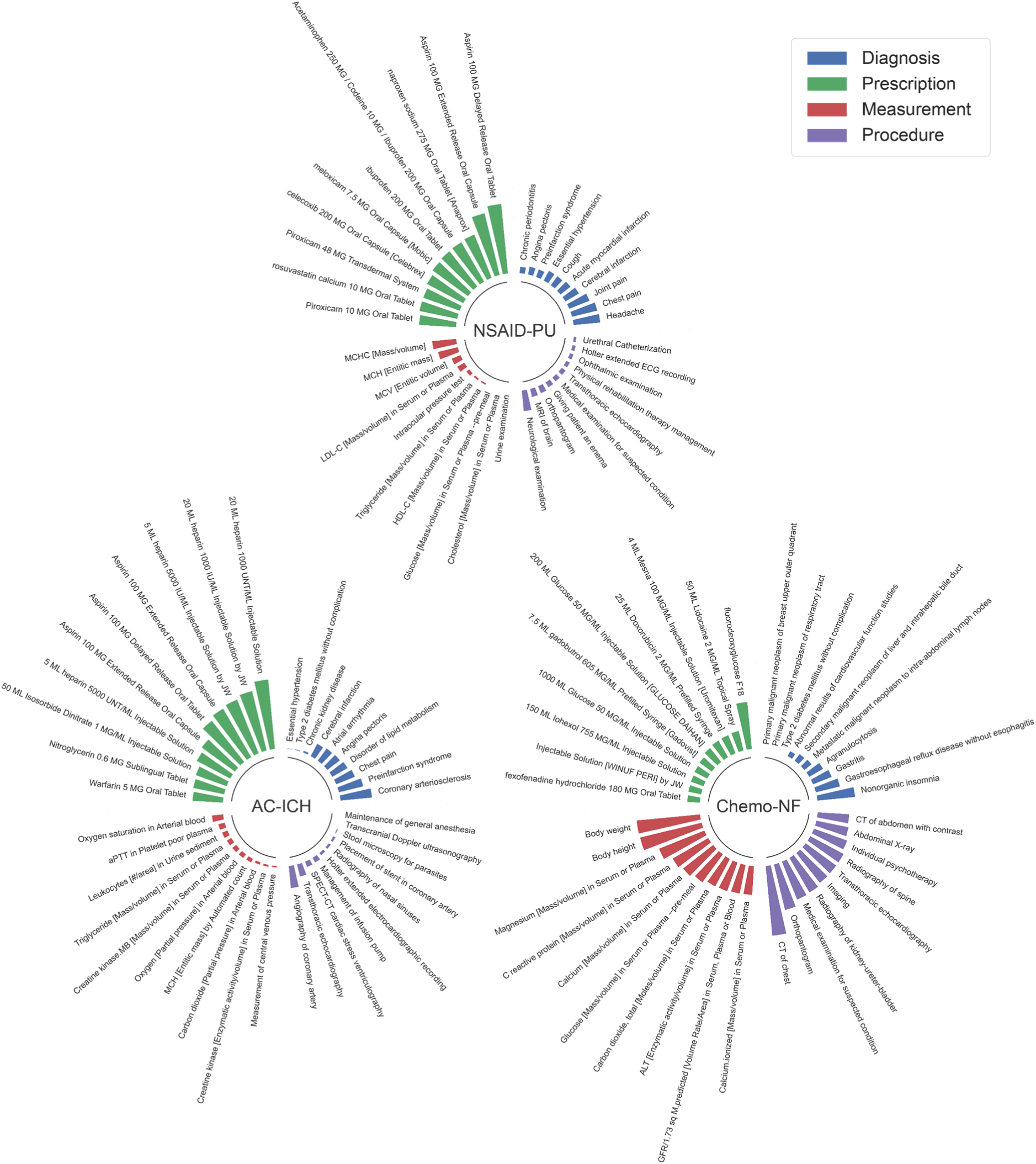
Feature importance of models finetuned with the external dataset. Top 10 important features in each domain. The feature importance calculation process was the same as that used for the internal dataset.

For the AC-ICH cohort in the internal validation, the models perceived aspirin and heparin prescription patterns as important for ICH prediction. The most important diagnosis was skin lesion, which might be related to bruising or petechiae caused by AC, and clinicians might have simply coded the condition as a skin lesion rather than its own name.^28,29^ Headache and dizziness, which are the main symptoms of ICH, were also important.^30,31^ The second most important diagnosis, hypercholesterolemia, is closely related to cardiovascular disease and can be associated with ICH.^32,33^ For measurements, lipid profile (triglyceride, HDL-cholesterol, LDL-cholesterol), which is a direct indicator for assessing hypercholesterolemia, and prothrombin time (PT) and fibrinogen, which are primarily focused on ICH patients,^34–36^ were included in the top 10 most important measurements. The models also closely observed procedures for diagnosis of cerebrovascular diseases, such as computed tomography (CT) of brain and transcranial Doppler (TCD) ultrasonography. The feature importance calculation result of the internal validation well-reflected prior knowledge regarding AC and ICH, but an unexpected feature (gastroesophageal reflux disease (GERD)) still existed. In the external validation, there were many features regarding heart diseases: coronary arteriosclerosis, preinfarction syndrome, angiography of coronary artery, and transthoracic echocardiography. Compared to the internal validation, the model paid less attention to the measurement, but several factors associated with heart disease (aPTT and creatine kinase-MB)^37,38^ were included in the top 10. Accordingly, in this cohort, the patients with heart disease might have developed cerebrovascular disease as well. For the CT-NF cohort in the internal validation, several drugs for chemotherapy (docetaxel, cisplatin, cyclophosphamide, rituximab, carboplatin, and paclitaxel) were included. Netupitant and palonosetron are drugs for preventing chemotherapy-induced nausea and vomiting.^39,40^ Patients who had experienced breast cancer were more likely to suffer from NF in this cohort. Chronic type B viral hepatitis is closely related to hepatocellular carcinoma. For measurement, carcinoembryonic antigen, which is an indicator for cancer diagnosis and monitoring,^41^ and inflammation-related features such as C reactive protein (CRP) and erythrocyte sedimentation rate (ESR) were included in the top 10 most important measurements. Central venous catheter (CVC) and PET CT FDG, which are closely associated with cancer, were important procedures. Screening colonoscopy is essential for the detection of colon cancer, and Nulytely is used for bowel cleansing prior to colonoscopy.^42,43^ Several types of CT and ultrasonography guided puncture and spiration of abdomen, which are procedures for the detection of cancer, were also included. Iron stain method, which is usually executed for the detection of hematologic malignancy, one of the highest-risk cancers for NF, was another important procedure.^44,45^ In the external validation, fluorodeoxyglucose (FDG) F18, gadobutrol, and iohexol, which are contrast medium drugs for cancer detection, were focused on more than chemotherapy drugs. Contrary to the internal validation, it is notable that agranulocytosis, which usually precedes NF,^46^ was included in the top 10.

In addition to cohort-level interpretation, patient-level interpretation is also available. We reported a sample of patient-level interpretation in the Chemo-NF cohort (Supplementary Fig. 2). Patient-level interpretation allows clinicians to understand which records and timepoints were important for predicting ADE for each patient.

## Discussion

We have proposed, externally validated, and qualitatively analyzed EHR pretraining model pretrained on CDM-based EHR data with DE. CDM schema consists of several domains, and we utilized data from four domains, including diagnosis, measurement, prescription, and procedure. We introduced DE in EHR pretraining model to let the model focus only on a necessary domain, and DE improved the performance of almost all the ADE prediction tasks. For external validation, the type II pretrained model outperformed all the other models, demonstrating the effectiveness of the model initially pretrained by a different dataset. The qualitative analysis with the importance of features at the cohort and patient levels was available, and the attention scores of each finetuned model well-reflected the relevant background clinical knowledge and the characteristics of the corresponding cohort.

We validated that the initially pretrained model works with another dataset. In the external validation, the type I pretrained model outperformed initially randomized models in all cohorts, even though they were never aware of the external dataset. In addition, the type II pretrained model achieved significantly improved performance with only ten training epochs. Given that only around 7,400 codes were common between the two hospitals, which had 26,417 and 22,551 codes, respectively, it is noteworthy that the model pretrained with numerous unknown codes remained effective. Additional pretraining successfully resolved the disparity of different datasets and improved performance. The initially pretrained model was also effective in terms of time efficiency. It took around six hours to pretrain the external dataset for one iteration with our single GPU. Considering we spent around a week for 100 iterations with four multiple GPUs, the ability to develop a strong pretrained model with fewer epochs can be especially attractive to smaller institutions with limited computational resources.

Hospital data has clear pros and cons for EHR pretraining: The upside is that the patient data generated within the hospital is real-time and of very high quality, and the downside is that it is very difficult to access data outside the hospital. Survey data can supplement historical medical records to some extent, but it is not enough. Accordingly, it is difficult to predict diseases, mortality, and readmission, which are very affected by data outside the hospital, using in-hospital data.^9–12,47^ To maximize the advantages of hospital data, we applied the EHR pretraining model to ADE prediction for inpatients. A pretraining model can learn changes before and after taking medication. Thus, the pretraining process included a deep understanding of drug reactions.

Compatibility is a significant advantage of CDM-based EHR pretraining model. OMOP-CDM is an open community data standard and is represented in more than 19 countries, with more than 200 million patient records, and more than 2,500 collaborators.^17,48^ The model pretrained from one hospital can be applied to, enhanced by, and validated by many other global hospital datasets.^15,16^ In addition, there is a national institution called Health Insurance Review and Assessment Service (HIRA) in Korea that has claims data converted to OMOP CDM format, and claims data can complement the lack of patient follow-up in hospital data.^49–51^ A large scale model through federated learning algorithms is also available.^52^ Several studies have already conducted multi-institutional data analysis via the federated learning framework for OMOP CDM data.^53,54^ CDM-based EHR pretraining model trained by data at the regional or national level can serve as a foundation model for various tasks in addition to in-hospital tasks such as ADE prediction.

This study has several limitations. First, we defined cohorts very roughly, even though several critical exclusion criteria might exist for proper cohort analysis. This approach was to demonstrate the effectiveness of EHR pretraining models in predicting various ADEs. For future work, we suggest conducting cohort studies with more rigorously defined cohort criteria and detailed analysis using our proposed methods. Second, there were irrelevant important features for each cohort. For example, speech audiometry, which is extraneous with all the ADE in this study, was found in the NSAID-PU and AC-ICH cohorts. It might be associated with overfitting, black box, and age, as PU and ICH usually occur in aged patients, but further investigation is needed. Additionally, for future work, we suggest leveraging large language models (LLMs) to provide well-established explanations with identified known risk factors for each patient at ADE risk, making the results more interpretable for clinicians. Third, the percentage of shared medical codes between the internal and external datasets was too low. Although this discrepancy demonstrated the effectiveness of the model pretrained with numerous unknown codes, alternatives to mitigate the inconsistency of vocabulary should be investigated more. Fourth, our model was only trained by structured EHR data from a single nation. For future work, we suggest utilizing and combining various types of unstructured medical data, as well as conducting external validation using global datasets.

In conclusion, we have demonstrated the potential of CDM-based EHR pretraining model as a foundation model through its prediction performance, interpretability, and compatibility. The adoption of domain embedding was effective in almost all cases, simplifying the pretraining procedures and improving comprehension of a medical context. The model interpretability for each cohort was consistent with several prior studies and clinical knowledge. Enhanced code systems to mitigate vocabulary inconsistency and CDM-based EHR pretraining model combined with unstructured data types are suggested for future works.

## Data Availability

All data produced in the present study are available upon reasonable request to the authors.

## Data availability

The datasets of SNUH and AUMC are not publicly available due to patient privacy and are available from the corresponding author on reasonable request and IRB approvals.

## Code availability

Source code for the experiments is publicly available at https://github.com/kicarussays/CDM-BERT.

## Author contributions

J.K., J.S.K., and K.K. conceptualized the study. J.K. conducted data curation, model development, and statistical analysis. K.K., M.G.K., and R.W.P. provided data and managed data security issues. J.H.L., T.K., C.C., and R.W.P. reviewed clinical validity of the study. J.K. wrote the original draft. J.S.K., J.H.L., and R.W.P. edited the draft.

