## Supplementary materials for "Pretrained Patient Trajectories for Adverse Drug Event Prediction Using Common Data Model-based Electronic Health Records"

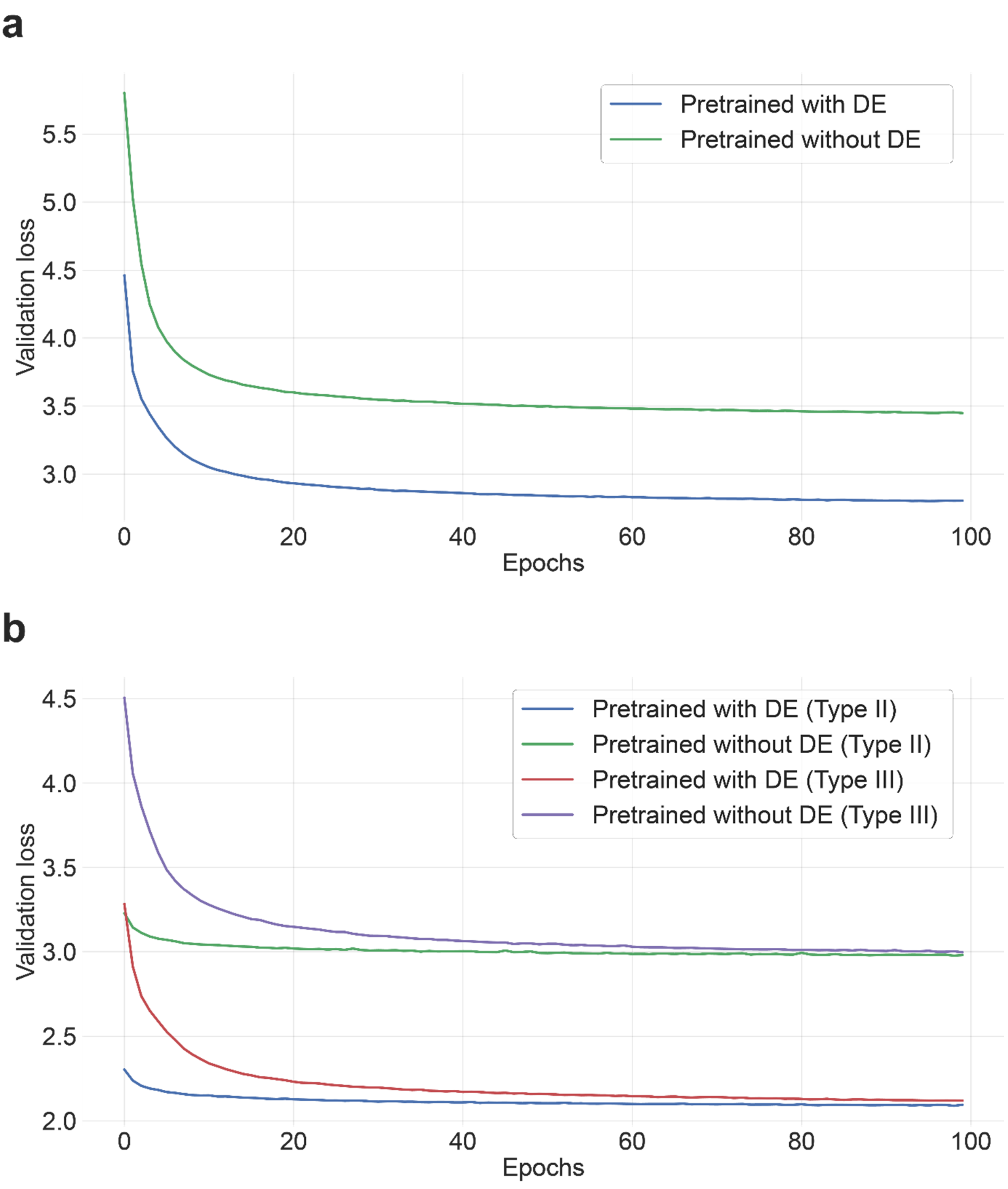


**Supplementary Fig. 1. Loss graphs of all pretraining tasks. a,** internal dataset**. b,** external dataset. The type I pretrained model was omitted since it was not additionally pretrained. For all pretraining tasks, the loss was much lower with the adoption of domain embedding. For the type II and III pretrained models, the validation loss was much lower when the models were additionally pretrained from the model initially pretrained by the other dataset (Type II).

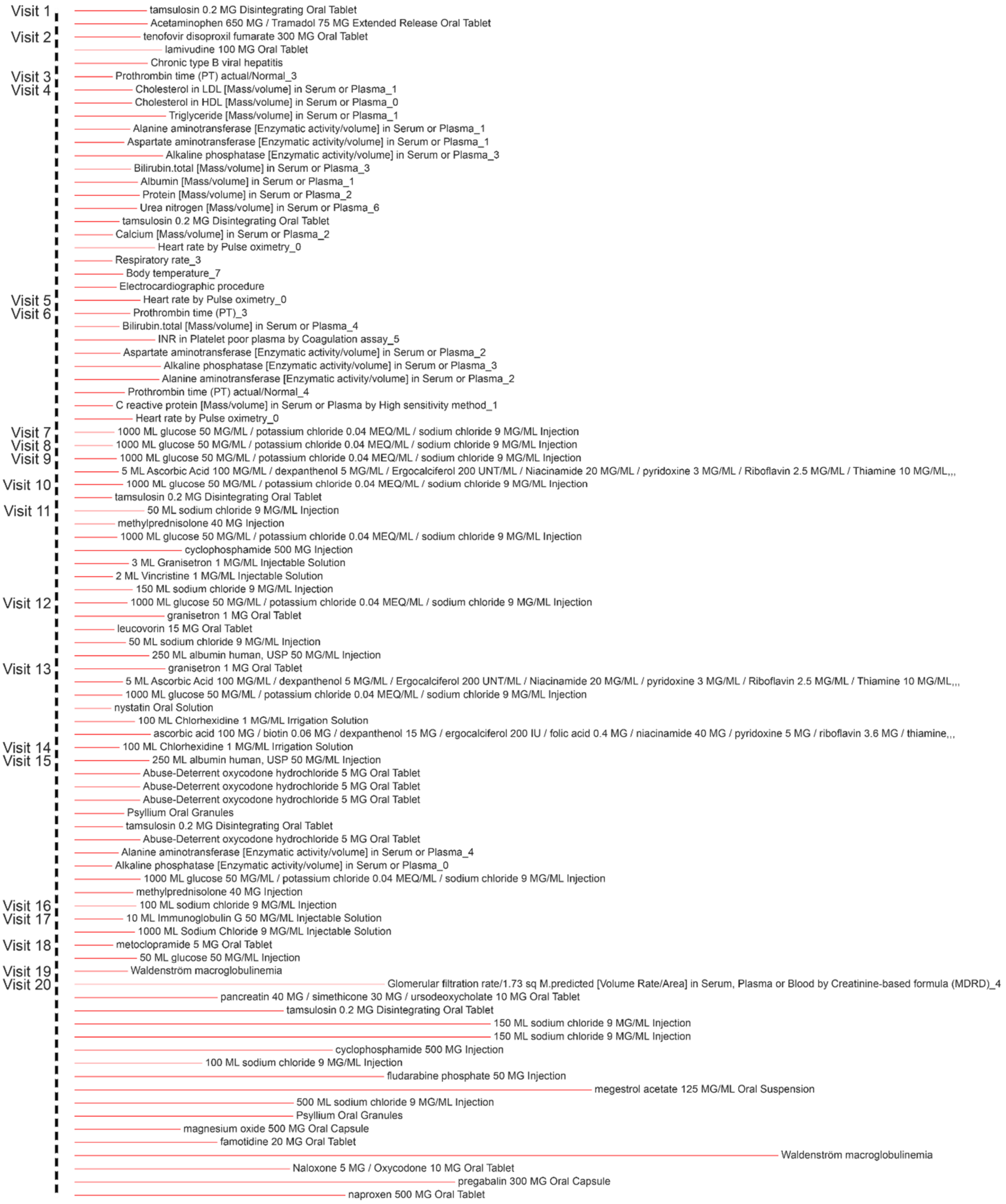


**Supplementary Fig. 2. Patient-level feature importance sample (Chemo-NF).** For individualized analysis, we simply summed all rows of an attention matrix as the sum of each row is 1, then extracted the top 10% of tokens with the highest values. This process was to enhance readability because most trajectories had hundreds of tokens. This allows clinicians to understand which records and timepoints were important in predicting adverse drug events for each patient.

**Supplementary Table 1. Drugs included in each drug group.**

| Drug groups | Drugs |
|---|---|
| NSAID | Aspirin, Diclofenac, Aceclofenac, Indomethacin, Ibuprofen, Naproxen, Celecoxib, Flurbiprofen, Fenoprofen, Ketoprofen, Loxoprofen, Oxaprozine, Piroxicam, Meloxicam |
| Anticoagulant | Warfarin, Heparin, NOAC (Rivaroxaban, Apixaban, Edoxaban, Dabigatran), Enoxaparin, Dalteparin |
| Alkylating agent | Cyclophosphamide, Bendamustine, Melphalan, Cisplatin, Carboplatin, Oxaliplatin, Busulfan, Dacarbazine, Temozolomide |
| Antimetabolite | Fluorouracil, Capecitabine, Doxifluridine, Tegafur, Azacitidine, Decitabine, Enocitabine, Methotrexate, Pemetrexed, Pralatrexate, Cladribine, Fludarabine, Mercaptopurine |
| Topoisomerase inhibitor | Doxorubicin, Daunorubicin, Epirubicin, Idarubicin, Mitoxantrone, Etoposide, Irinotecan, Topotecan |
| Microtubule targeting agent | Cabazitaxel, Paclitaxel, Docetaxel, Vinblastine, Vincristine, Vinorelbine |

NSAIDs, nonsteroidal anti-inflammatory drugs; NOAC, non-vitamin K antagonist oral anticoagulant

**Supplementary Table 2. Concept IDs for each adverse drug event. All concept IDs belong to SNOMED-CT.**

| Adverse drug events | Concept IDs |
|---|---|
| Peptic ulcer (PU) | 4146517, 36683388, 4194543, 45757062, 45757242, 198798, 4006994, 434509, 4046500, 760961, 4131615, 4130998, 4028242, 4134146, 4027729, 4027663, 4059178, 4066036, 4087594, 4057060, 195306, 4057687, 4057953, 4049350, 197418, 4341234, 4341240, 4341242, 4340787, 4341078, 4112610, 4115272, 4156639, 4101104, 4101870, 441067, 4180023, 4147683, 4143871, 25844, 4150681, 4138962, 443386, 4178966, 4289830, 441225, 4291028, 4212919, 4265600, 4222896, 4256775, 4215084, 4212693, 4177387, 4330087, 4206524, 4163865, 4164920, 4169592, 4174044, 4173408, 4196964, 4198381, 442270, 4200399, 4204555, 4209746, 4211001, 26441, 4263697, 4247008, 4265479, 433515, 4274491, 4271696, 4279470, 4280942, 4195231, 37396011, 433516, 36717179, 36715928, 36715929, 37110314, 4248429, 4251486, 42538071, 4294973, 4296611, 4217947, 4336230, 4338225, 4232181, 4231580, 4235753, 4242191, 436062, 4243739, 435752, 4313355, 4317413, 4319441, 4318534, 434085 |
| Intracranial hemorrhage (ICH) | 42872427, 4306943, 4112018, 4017105, 439040, 37394466, 4130539, 439847, 43530674, 4108952, 4111708, 4110185, 4110186, 4111709, 4111720, 4111721, 4071589, 4049659, 4014781, 4017107, 260841, 432923, 4045744, 4045745, 4046365, 443752, 4136546, 4134162, 4148906, 376713, 4077828, 4078446, 4077200, 4077958, 4077959, 4078448, 4106058, 42535424, 42535425, 42539269, 43530727, 42535426, 4199890, 4121664, 4159150, 4159151, 436430, 4176148, 40492969, 42873157, 4176892, 441709, 435960, 436519, 37016924, 36716572, 36716581, 36716627, 36716862, 4249574, 4218781, 42537643, 4326561, 4299377, 3654885, 3654892, 3654894, 4318408, 4319328 |
| Neutropenic fever (NF) | 4250734 |

**Supplementary Table 3. Token counts for each domain in each dataset.**

| | Internal dataset | External dataset | Shared | Aggregated |
|---|---|---|---|---|
| Diagnosis | 12,913 | 10,420 | 5,548 | 17,785 |
| Prescription | 6,152 | 6,332 | 271 | 12,213 |
| Measurement | 2,462 | 2,857 | 1,064 | 4,255 |
| Procedure | 4,883 | 2,935 | 542 | 7,276 |
| Total | 26,410 + 5 special tokens + 2 gender tokens | 22,544 + 5 special tokens + 2 gender tokens | 7,425 + 5 special tokens + 2 gender tokens | 41529 + 5 special tokens + 2 gender tokens |

**Supplementary Table 4. Patient demographics of each cohort.**

| | Prediction timepoint | Case group | | | Control group | | | Incidence ratio |
|---|---|---|---|---|---|---|---|---|
| | | Count | Age | Male sex | Count | Age | Male sex | |
| SNUH (internal) | | | | | | | | |
| NSAID (PU) | 2 weeks | 1880 | 64.89±12.45 | 40.48% | 264925 | 55.11±15.99 | 58.32% | 0.70% |
| | 4 weeks | 2345 | 64.92±12.36 | 41.07% | 264925 | 55.11±15.99 | 58.32% | 0.88% |
| | 8 weeks | 2880 | 65.08±12.20 | 41.81% | 264925 | 55.11±15.99 | 58.32% | 1.08% |
| | 12 weeks | 3281 | 64.94±12.13 | 41.63% | 264925 | 55.11±15.99 | 58.32% | 1.22% |
| AC (ICH) | 2 weeks | 883 | 63.90±14.45 | 47.45% | 155586 | 61.28±14.33 | 46.26% | 0.56% |
| | 4 weeks | 1073 | 64.68±13.87 | 47.34% | 155586 | 61.28±14.33 | 46.26% | 0.68% |
| | 8 weeks | 1262 | 65.50±13.54 | 47.54% | 155586 | 61.28±14.33 | 46.26% | 0.80% |
| | 12 weeks | 1396 | 66.04±13.50 | 46.70% | 155586 | 61.28±14.33 | 46.26% | 0.89% |
| Chemo (NF) | 2 weeks | 2064 | 56.62±14.17 | 62.35% | 82803 | 58.14±12.98 | 50.11% | 2.43% |
| | 4 weeks | 2519 | 56.87±13.97 | 61.73% | 82803 | 58.14±12.98 | 50.11% | 2.95% |
| | 8 weeks | 2354 | 57.11±13.88 | 61.17% | 82803 | 58.14±12.98 | 50.11% | 2.76% |
| | 12 weeks | 2162 | 57.62±13.72 | 59.81% | 82803 | 58.14±12.98 | 50.11% | 2.54% |
| AUMC (external) | | | | | | | | |
| NSAID (PU) | 2 weeks | 2451 | 60.22±13.55 | 50.92% | 176223 | 54.47±16.94 | 51.04% | 1.37% |
| | 4 weeks | 3043 | 60.04±13.57 | 50.51% | 176223 | 54.47±16.94 | 51.04% | 1.70% |
| | 8 weeks | 3587 | 60.17±13.38 | 49.54% | 176223 | 54.47±16.94 | 51.04% | 1.99% |
| | 12 weeks | 3903 | 60.15±13.42 | 48.94% | 176223 | 54.47±16.94 | 51.04% | 2.17% |
| AC (ICH) | 2 weeks | 1099 | 58.99±14.34 | 45.13% | 117407 | 60.24±15.57 | 45.53% | 0.93% |
| | 4 weeks | 1228 | 59.58±14.50 | 44.95% | 117407 | 60.24±15.57 | 45.53% | 1.04% |
| | 8 weeks | 1397 | 60.53±14.39 | 44.52% | 117407 | 60.24±15.57 | 45.53% | 1.18% |
| | 12 weeks | 1506 | 61.14±14.38 | 44.56% | 117407 | 60.24±15.57 | 45.53% | 1.27% |
| Chemo (NF) | 2 weeks | 364 | 56.94±13.27 | 71.15% | 34063 | 57.07±13.63 | 50.70% | 1.06% |
| | 4 weeks | 406 | 57.69±13.30 | 68.97% | 34063 | 57.07±13.63 | 50.70% | 1.18% |
| | 8 weeks | 384 | 57.72±13.01 | 65.62% | 34063 | 57.07±13.63 | 50.70% | 1.11% |
| | 12 weeks | 346 | 57.81±13.07 | 62.43% | 34063 | 57.07±13.63 | 50.70% | 1.01% |

SNUH, Seoul National University Hospital; AUMC, Ajou University Medical Center; NSAID, nonsteroidal anti-inflammatory drugs; PU, peptic ulcer; AC, anticoagulants; ICH, intracranial hemorrhage; Chemo, chemotherapy; NF, neutropenic fever

**Supplementary Table 5. Internal validations of finetuned models with different prediction timepoint (two weeks before the occurrence of adverse events).**

| | AUROC | AUPRC | Sensitivity | Specificity | Precision | F1-score |
|---|---|---|---|---|---|---|
| *Internal validation (SNUH)* | | | | | | |
| NSAID (PU) | | | | | | |
| Randomized without DE | 0.834 (0.814-0.853) | 0.034 (0.014-0.053) | 0.794 (0.791-0.798) | 0.736 (0.732-0.739) | 0.019 (0.018-0.020) | 0.037 (0.036-0.039) |
| Randomized with DE | 0.834 (0.815-0.853) | 0.034 (0.015-0.053) | 0.791 (0.788-0.795) | 0.740 (0.736-0.743) | 0.019 (0.018-0.021) | 0.038 (0.036-0.040) |
| Pretrained without DE | 0.948 (0.938-0.958) | 0.279 (0.269-0.288) | 0.896 (0.893-0.898) | 0.855 (0.852-0.858) | 0.039 (0.037-0.040) | 0.074 (0.072-0.076) |
| Pretrained with DE | **0.982 (0.978-0.987)** | **0.604 (0.599-0.608)** | **0.936 (0.934-0.938)** | **0.921 (0.919-0.923)** | **0.072 (0.070-0.074)** | **0.133 (0.130-0.136)** |
| AC (ICH) | | | | | | |
| Randomized without DE | 0.706 (0.668-0.743) | 0.015 (0.000-0.743) | 0.725 (0.720-0.730) | 0.590 (0.585-0.596) | 0.011 (0.009-0.012) | 0.021 (0.019-0.022) |
| Randomized with DE | 0.705 (0.668-0.743) | 0.015 (0.000-0.743) | 0.725 (0.720-0.730) | 0.590 (0.585-0.595) | 0.011 (0.009-0.012) | 0.021 (0.019-0.022) |
| Pretrained without DE | 0.717 (0.678-0.755) | 0.027 (0.000-0.755) | 0.598 (0.592-0.603) | 0.734 (0.729-0.739) | 0.013 (0.012-0.015) | 0.026 (0.024-0.028) |
| Pretrained with DE | **0.826 (0.793-0.858)** | **0.174 (0.141-0.207)** | **0.757 (0.752-0.761)** | **0.744 (0.739-0.748)** | **0.017 (0.016-0.019)** | **0.034 (0.032-0.036)** |
| CT (NF) | | | | | | |
| Randomized without DE | 0.831 (0.813-0.848) | 0.116 (0.099-0.134) | 0.744 (0.737-0.750) | 0.779 (0.773-0.785) | 0.078 (0.074-0.082) | 0.140 (0.135-0.146) |
| Randomized with DE | 0.831 (0.813-0.848) | 0.116 (0.099-0.134) | 0.744 (0.737-0.750) | 0.779 (0.773-0.785) | 0.077 (0.073-0.082) | 0.140 (0.135-0.146) |
| Pretrained without DE | 0.976 (0.971-0.982) | 0.747 (0.742-0.753) | 0.918 (0.914-0.922) | 0.917 (0.913-0.921) | 0.216 (0.210-0.222) | 0.350 (0.342-0.357) |
| Pretrained with DE | **0.992 (0.989-0.995)** | **0.902 (0.899-0.905)** | **0.957 (0.953-0.959)** | **0.949 (0.946-0.952)** | **0.319 (0.312-0.326)** | **0.479 (0.471-0.486)** |

Bold indicates the best. Sensitivity, specificity, precision, and F1-score were calculated using Youden's index. Confidence intervals (CIs) of AUROC and AUPRC were calculated using DeLong's method. CIs of sensitivity, specificity, precision, and F1-score were calculated using Wilson's method. NSAID, nonsteroidal anti-inflammatory drug; PU, peptic ulcer; AC, anticoagulant; ICH, intracranial hemorrhage; CT, chemotherapy; NF, neutropenic fever; AUROC, area under the receiver operating characteristic curve; AUPRC, area under the precision-recall curve

**Supplementary Table 6. External validations of finetuned models with different prediction timepoint (two weeks before the occurrence of adverse events).**

| | AUROC | AUPRC | Sensitivity | Specificity | Precision | F1-score |
|---|---|---|---|---|---|---|
| *External validation (AUMC)* | | | | | | |
| NSAID (PU) | | | | | | |
| Randomized without DE | 0.816 (0.799-0.834) | 0.056 (0.038-0.073) | 0.837 (0.833-0.841) | 0.666 (0.661-0.671) | 0.032 (0.030-0.034) | 0.062 (0.059-0.064) |
| Randomized with DE | 0.816 (0.799-0.834) | 0.056 (0.038-0.073) | 0.846 (0.842-0.850) | 0.658 (0.653-0.663) | 0.032 (0.030-0.034) | 0.061 (0.059-0.064) |
| Pretrained without DE (Type II) | 0.908 (0.894-0.921) | 0.262 (0.248-0.276) | 0.842 (0.838-0.845) | 0.820 (0.816-0.824) | 0.058 (0.056-0.061) | 0.109 (0.106-0.112) |
| Pretrained with DE (Type II) | **0.967 (0.959-0.975)** | **0.674 (0.666-0.682)** | **0.929 (0.927-0.932)** | **0.885 (0.881-0.888)** | **0.096 (0.093-0.099)** | **0.175 (0.171-0.178)** |
| AC (ICH) | | | | | | |
| Randomized without DE | 0.756 (0.720-0.792) | 0.078 (0.042-0.114) | 0.676 (0.670-0.682) | 0.739 (0.734-0.745) | 0.022 (0.020-0.024) | 0.043 (0.040-0.045) |
| Randomized with DE | 0.748 (0.712-0.784) | 0.076 (0.039-0.112) | 0.628 (0.622-0.634) | 0.768 (0.762-0.773) | 0.023 (0.021-0.025) | 0.044 (0.042-0.047) |
| Pretrained without DE (Type II) | 0.947 (0.930-0.964) | 0.669 (0.652-0.686) | 0.821 (0.816-0.826) | **0.938 (0.935-0.941)** | **0.103 (0.100-0.107)** | **0.184 (0.179-0.189)** |
| Pretrained with DE (Type II) | **0.972 (0.960-0.983)** | **0.731 (0.720-0.742)** | **0.903 (0.900-0.907)** | 0.927 (0.923-0.930) | 0.097 (0.093-0.101) | 0.175 (0.170-0.180) |
| CT (NF) | | | | | | |
| Randomized without DE | 0.914 (0.881-0.947) | 0.262 (0.229-0.295) | 0.859 (0.851-0.867) | 0.865 (0.857-0.873) | 0.062 (0.057-0.068) | 0.116 (0.109-0.124) |
| Randomized with DE | 0.914 (0.881-0.947) | 0.263 (0.230-0.296) | 0.859 (0.851-0.867) | 0.865 (0.856-0.872) | 0.062 (0.057-0.068) | 0.116 (0.108-0.123) |
| Pretrained without DE (Type II) | 0.949 (0.930-0.968) | 0.366 (0.347-0.385) | **0.958 (0.953-0.962)** | 0.831 (0.822-0.840) | 0.056 (0.051-0.061) | 0.105 (0.098-0.113) |
| Pretrained with DE (Type II) | **0.972 (0.959-0.985)** | **0.594 (0.581-0.607)** | 0.915 (0.909-0.922) | **0.896 (0.889-0.903)** | **0.084 (0.078-0.091)** | **0.154 (0.146-0.163)** |

Bold indicates the best. Sensitivity, specificity, precision, and F1-score were calculated using Youden's index. Confidence intervals (CIs) of AUROC and AUPRC were calculated using DeLong's method. CIs of sensitivity, specificity, precision, and F1-score were calculated using Wilson's method. NSAID, nonsteroidal anti-inflammatory drug; PU, peptic ulcer; AC, anticoagulant; ICH, intracranial hemorrhage; CT, chemotherapy; NF, neutropenic fever; AUROC, area under the receiver operating characteristic curve; AUPRC, area under the precision-recall curve

**Supplementary Table 7. Internal validations of finetuned models with different prediction timepoint (eight weeks before the occurrence of adverse events).**

| | AUROC | AUPRC | Sensitivity | Specificity | Precision | F1-score |
|---|---|---|---|---|---|---|
| *Internal validation (SNUH)* | | | | | | |
| NSAID (PU) | | | | | | |
| Randomized without DE | 0.860 (0.846-0.874) | 0.062 (0.048-0.076) | 0.792 (0.788-0.795) | 0.787 (0.784-0.791) | 0.038 (0.037-0.040) | 0.073 (0.071-0.075) |
| Randomized with DE | 0.860 (0.846-0.874) | 0.062 (0.048-0.076) | 0.792 (0.788-0.795) | 0.787 (0.784-0.790) | 0.038 (0.037-0.040) | 0.073 (0.071-0.075) |
| Pretrained without DE | 0.965 (0.958-0.971) | 0.532 (0.526-0.538) | 0.873 (0.870-0.876) | 0.925 (0.923-0.928) | 0.111 (0.109-0.114) | 0.197 (0.194-0.201) |
| Pretrained with DE | **0.983 (0.978-0.987)** | **0.672 (0.668-0.677)** | **0.937 (0.934-0.939)** | **0.945 (0.943-0.947)** | **0.154 (0.151-0.157)** | **0.265 (0.261-0.268)** |
| AC (ICH) | | | | | | |
| Randomized without DE | 0.741 (0.710-0.771) | 0.026 (0.000-0.771) | 0.698 (0.693-0.704) | 0.691 (0.686-0.696) | 0.019 (0.017-0.020) | 0.036 (0.034-0.038) |
| Randomized with DE | 0.741 (0.710-0.771) | 0.026 (0.000-0.771) | 0.698 (0.693-0.704) | 0.691 (0.686-0.696) | 0.018 (0.017-0.020) | 0.036 (0.034-0.038) |
| Pretrained without DE | 0.885 (0.865-0.904) | 0.120 (0.100-0.139) | 0.828 (0.824-0.832) | 0.802 (0.797-0.806) | 0.034 (0.032-0.036) | 0.065 (0.062-0.067) |
| Pretrained with DE | **0.953 (0.940-0.965)** | **0.395 (0.383-0.408)** | **0.870 (0.866-0.874)** | **0.896 (0.892-0.899)** | **0.065 (0.062-0.068)** | **0.121 (0.117-0.124)** |
| CT (NF) | | | | | | |
| Randomized without DE | 0.830 (0.812-0.847) | 0.136 (0.119-0.153) | 0.832 (0.827-0.838) | 0.679 (0.672-0.686) | 0.065 (0.062-0.069) | 0.121 (0.116-0.126) |
| Randomized with DE | 0.830 (0.813-0.847) | 0.136 (0.119-0.153) | 0.830 (0.824-0.836) | 0.683 (0.676-0.690) | 0.066 (0.062-0.070) | 0.122 (0.117-0.127) |
| Pretrained without DE | 0.981 (0.976-0.986) | 0.785 (0.780-0.789) | **0.964 (0.961-0.967)** | 0.895 (0.891-0.900) | 0.199 (0.193-0.205) | 0.329 (0.322-0.336) |
| Pretrained with DE | **0.984 (0.979-0.989)** | **0.865 (0.859-0.870)** | 0.924 (0.920-0.928) | **0.947 (0.944-0.951)** | **0.320 (0.313-0.327)** | **0.476 (0.468-0.483)** |

Bold indicates the best. Sensitivity, specificity, precision, and F1-score were calculated using Youden's index. Confidence intervals (CIs) of AUROC and AUPRC were calculated using DeLong's method. CIs of sensitivity, specificity, precision, and F1-score were calculated using Wilson's method. NSAID, nonsteroidal anti-inflammatory drug; PU, peptic ulcer; AC, anticoagulant; ICH, intracranial hemorrhage; CT, chemotherapy; NF, neutropenic fever; AUROC, area under the receiver operating characteristic curve; AUPRC, area under the precision-recall curve

**Supplementary Table 8. External validations of finetuned models with different prediction timepoint (eight weeks before the occurrence of adverse events).**

| | AUROC | AUPRC | Sensitivity | Specificity | Precision | F1-score |
|---|---|---|---|---|---|---|
| *External validation (AUMC)* | | | | | | |
| NSAID (PU) | | | | | | |
| Randomized without DE | 0.825 (0.811-0.839) | 0.085 (0.070-0.099) | 0.845 (0.842-0.849) | 0.681 (0.676-0.686) | 0.049 (0.047-0.051) | 0.093 (0.090-0.096) |
| Randomized with DE | 0.825 (0.811-0.839) | 0.085 (0.070-0.099) | 0.845 (0.842-0.849) | 0.681 (0.676-0.686) | 0.049 (0.047-0.051) | 0.093 (0.090-0.096) |
| Pretrained without DE (Type II) | 0.912 (0.898-0.926) | 0.419 (0.405-0.434) | 0.879 (0.876-0.882) | 0.850 (0.846-0.854) | 0.102 (0.099-0.105) | 0.183 (0.179-0.187) |
| Pretrained with DE (Type II) | **0.980 (0.975-0.985)** | **0.764 (0.759-0.770)** | **0.958 (0.956-0.960)** | **0.905 (0.902-0.908)** | **0.164 (0.160-0.167)** | **0.279 (0.275-0.284)** |
| AC (ICH) | | | | | | |
| Randomized without DE | 0.795 (0.768-0.823) | 0.080 (0.052-0.107) | 0.739 (0.733-0.744) | 0.732 (0.727-0.738) | 0.031 (0.028-0.033) | 0.059 (0.056-0.062) |
| Randomized with DE | 0.795 (0.768-0.823) | 0.080 (0.052-0.107) | 0.699 (0.693-0.704) | 0.774 (0.769-0.779) | 0.034 (0.032-0.037) | 0.065 (0.062-0.068) |
| Pretrained without DE (Type II) | 0.937 (0.921-0.953) | 0.541 (0.525-0.557) | 0.875 (0.871-0.879) | 0.875 (0.871-0.879) | 0.074 (0.071-0.078) | 0.137 (0.133-0.141) |
| Pretrained with DE (Type II) | **0.973 (0.963-0.983)** | **0.709 (0.700-0.719)** | **0.908 (0.904-0.912)** | **0.935 (0.932-0.938)** | **0.137 (0.133-0.142)** | **0.238 (0.233-0.244)** |
| CT (NF) | | | | | | |
| Randomized without DE | 0.881 (0.843-0.918) | 0.169 (0.131-0.206) | 0.846 (0.837-0.854) | 0.791 (0.781-0.800) | 0.044 (0.040-0.049) | 0.084 (0.078-0.091) |
| Randomized with DE | 0.881 (0.843-0.918) | 0.169 (0.131-0.206) | 0.846 (0.837-0.854) | 0.791 (0.781-0.800) | 0.044 (0.040-0.049) | 0.084 (0.078-0.091) |
| Pretrained without DE (Type II) | 0.906 (0.879-0.933) | 0.266 (0.239-0.294) | 0.859 (0.851-0.867) | **0.815 (0.806-0.824)** | **0.051 (0.046-0.056)** | **0.095 (0.089-0.103)** |
| Pretrained with DE (Type II) | **0.920 (0.892-0.947)** | **0.322 (0.294-0.350)** | **0.962 (0.957-0.966)** | 0.742 (0.732-0.752) | 0.041 (0.036-0.046) | 0.078 (0.072-0.085) |

Bold indicates the best. Sensitivity, specificity, precision, and F1-score were calculated using Youden's index. Confidence intervals (CIs) of AUROC and AUPRC were calculated using DeLong's method. CIs of sensitivity, specificity, precision, and F1-score were calculated using Wilson's method. NSAID, nonsteroidal anti-inflammatory drug; PU, peptic ulcer; AC, anticoagulant; ICH, intracranial hemorrhage; CT, chemotherapy; NF, neutropenic fever; AUROC, area under the receiver operating characteristic curve; AUPRC, area under the precision-recall curve

**Supplementary Table 9. Internal validations of finetuned models with different prediction timepoint (twelve weeks before the occurrence of adverse events).**

| | AUROC | AUPRC | Sensitivity | Specificity | Precision | F1-score |
|---|---|---|---|---|---|---|
| *Internal validation (SNUH)* | | | | | | |
| NSAID (PU) | | | | | | |
| Randomized without DE | 0.857 (0.843-0.871) | 0.080 (0.066-0.094) | 0.837 (0.834-0.840) | 0.741 (0.738-0.745) | 0.038 (0.037-0.040) | 0.074 (0.071-0.076) |
| Randomized with DE | 0.856 (0.842-0.870) | 0.081 (0.067-0.095) | 0.831 (0.828-0.834) | 0.744 (0.740-0.748) | 0.039 (0.037-0.040) | 0.074 (0.072-0.076) |
| Pretrained without DE | 0.972 (0.966-0.977) | 0.572 (0.567-0.578) | 0.934 (0.932-0.937) | 0.893 (0.890-0.895) | 0.097 (0.095-0.100) | 0.176 (0.173-0.180) |
| Pretrained with DE | **0.984 (0.980-0.989)** | **0.791 (0.786-0.796)** | **0.938 (0.935-0.940)** | **0.935 (0.933-0.937)** | **0.152 (0.149-0.155)** | **0.261 (0.258-0.265)** |
| AC (ICH) | | | | | | |
| Randomized without DE | 0.789 (0.763-0.815) | 0.040 (0.014-0.066) | 0.787 (0.782-0.791) | 0.660 (0.655-0.665) | 0.020 (0.019-0.022) | 0.039 (0.037-0.041) |
| Randomized with DE | 0.789 (0.763-0.815) | 0.040 (0.014-0.066) | 0.787 (0.782-0.791) | 0.661 (0.656-0.666) | 0.020 (0.019-0.022) | 0.039 (0.037-0.041) |
| Pretrained without DE | 0.924 (0.908-0.940) | 0.265 (0.249-0.281) | **0.913 (0.910-0.916)** | 0.803 (0.799-0.807) | 0.039 (0.037-0.041) | 0.075 (0.072-0.078) |
| Pretrained with DE | **0.978 (0.970-0.985)** | **0.676 (0.669-0.683)** | 0.903 (0.899-0.906) | **0.943 (0.941-0.946)** | **0.123 (0.120-0.127)** | **0.217 (0.212-0.221)** |
| CT (NF) | | | | | | |
| Randomized without DE | 0.832 (0.814-0.850) | 0.151 (0.133-0.169) | 0.815 (0.809-0.821) | 0.694 (0.687-0.701) | 0.063 (0.059-0.066) | 0.116 (0.112-0.121) |
| Randomized with DE | 0.832 (0.814-0.850) | 0.151 (0.133-0.169) | 0.817 (0.811-0.823) | 0.692 (0.685-0.699) | 0.062 (0.059-0.066) | 0.116 (0.111-0.121) |
| Pretrained without DE | 0.981 (0.976-0.986) | 0.780 (0.775-0.785) | 0.942 (0.939-0.946) | 0.923 (0.919-0.927) | 0.235 (0.229-0.242) | 0.377 (0.369-0.384) |
| Pretrained with DE | **0.993 (0.989-0.996)** | **0.942 (0.938-0.946)** | **0.947 (0.944-0.950)** | **0.987 (0.985-0.988)** | **0.642 (0.634-0.649)** | **0.765 (0.759-0.771)** |

Bold indicates the best. Sensitivity, specificity, precision, and F1-score were calculated using Youden's index. Confidence intervals (CIs) of AUROC and AUPRC were calculated using DeLong's method. CIs of sensitivity, specificity, precision, and F1-score were calculated using Wilson's method. NSAID, nonsteroidal anti-inflammatory drug; PU, peptic ulcer; AC, anticoagulant; ICH, intracranial hemorrhage; CT, chemotherapy; NF, neutropenic fever; AUROC, area under the receiver operating characteristic curve; AUPRC, area under the precision-recall curve

**Supplementary Table 10. External validations of finetuned models with different prediction timepoint (twelve weeks before the occurrence of adverse events).**

| | AUROC | AUPRC | Sensitivity | Specificity | Precision | F1-score |
|---|---|---|---|---|---|---|
| *External validation (AUMC)* | | | | | | |
| NSAID (PU) | | | | | | |
| Randomized without DE | 0.834 (0.821-0.847) | 0.101 (0.088-0.114) | 0.780 (0.775-0.784) | 0.729 (0.724-0.733) | 0.058 (0.056-0.061) | 0.108 (0.105-0.111) |
| Randomized with DE | 0.834 (0.821-0.847) | 0.101 (0.088-0.114) | 0.782 (0.778-0.787) | 0.728 (0.723-0.732) | 0.058 (0.056-0.061) | 0.108 (0.105-0.111) |
| Pretrained without DE (Type II) | 0.953 (0.946-0.960) | 0.531 (0.525-0.538) | 0.909 (0.906-0.912) | 0.875 (0.872-0.878) | 0.135 (0.132-0.139) | 0.235 (0.231-0.240) |
| Pretrained with DE (Type II) | **0.978 (0.973-0.983)** | **0.757 (0.752-0.762)** | **0.929 (0.926-0.931)** | **0.928 (0.925-0.931)** | **0.217 (0.213-0.221)** | **0.352 (0.347-0.357)** |
| AC (ICH) | | | | | | |
| Randomized without DE | 0.786 (0.758-0.813) | 0.087 (0.059-0.114) | 0.803 (0.798-0.808) | 0.670 (0.664-0.676) | 0.030 (0.028-0.032) | 0.057 (0.054-0.060) |
| Randomized with DE | 0.786 (0.759-0.814) | 0.087 (0.059-0.114) | 0.803 (0.798-0.808) | 0.670 (0.664-0.676) | 0.030 (0.028-0.032) | 0.057 (0.054-0.060) |
| Pretrained without DE (Type II) | 0.871 (0.850-0.892) | 0.394 (0.372-0.415) | 0.736 (0.730-0.741) | 0.831 (0.827-0.836) | 0.052 (0.049-0.055) | 0.097 (0.093-0.101) |
| Pretrained with DE (Type II) | **0.977 (0.969-0.986)** | **0.706 (0.698-0.715)** | **0.940 (0.937-0.943)** | **0.919 (0.916-0.923)** | **0.127 (0.123-0.132)** | **0.224 (0.219-0.230)** |
| CT (NF) | | | | | | |
| Randomized without DE | 0.873 (0.837-0.908) | 0.110 (0.074-0.146) | 0.824 (0.814-0.832) | 0.788 (0.778-0.797) | 0.037 (0.033-0.042) | 0.071 (0.065-0.078) |
| Randomized with DE | 0.872 (0.837-0.908) | 0.110 (0.074-0.146) | 0.824 (0.814-0.832) | 0.788 (0.779-0.798) | 0.037 (0.033-0.042) | 0.072 (0.066-0.078) |
| Pretrained without DE (Type II) | **0.919 (0.896-0.942)** | 0.198 (0.175-0.222) | **0.912 (0.905-0.918)** | 0.796 (0.786-0.805) | 0.043 (0.038-0.048) | 0.082 (0.075-0.088) |
| Pretrained with DE (Type II) | 0.913 (0.869-0.957) | **0.456 (0.412-0.500)** | 0.868 (0.859-0.875) | **0.849 (0.841-0.858)** | **0.054 (0.049-0.060)** | **0.102 (0.095-0.110)** |

Bold indicates the best. Sensitivity, specificity, precision, and F1-score were calculated using Youden's index. Confidence intervals (CIs) of AUROC and AUPRC were calculated using DeLong's method. CIs of sensitivity, specificity, precision, and F1-score were calculated using Wilson's method. NSAID, nonsteroidal anti-inflammatory drug; PU, peptic ulcer; AC, anticoagulant; ICH, intracranial hemorrhage; CT, chemotherapy; NF, neutropenic fever; AUROC, area under the receiver operating characteristic curve; AUPRC, area under the precision-recall curve
